# Modelling the effect of lockdown

**DOI:** 10.1101/2021.01.11.20248882

**Authors:** Hideto Kamei, Akihiro Sato

## Abstract

This note models the effect of the lockdown during the first wave of COVID-19. We use SEIR type of model with a certain time lag between infection and becoming infectious. Firstly we compare the timing of the change of the coefficient of infection, growth rate of confirmed cases corresponds to the change of mobility index, and secondly we assume the change of the coefficient of infection, activity index *β* (analogous to *R*_0_) and fit the parameter to reproduce the actual number of confirmed cases. Finally, we assume that the activity index *β* is proportional to the square of the mobility and fit the parameters. The curves in various cuontries fits reasonably well in any cases, but estimating *β* from various parameters (including temperature) remains as an important task.

## 2 Introduction

Due to the worldwide pandemic of COVID-19, many countries are trying to slow down the spreading of the virus through lockdown of the cities. At the time of writing this (2020/4/29) some has already achieved dramatic decrease in the number of patients, while most of them are still struggling, therefore it is crucial to evaluate the effect of the lockdown, equivalently, to estimate the time the lockdown is needed.

Therefore, we will try to model the dynamics for the number of patients, considering the effect of lockdown. We have roughly two different type of models which could explain the dynamics, one is ordinary SEIR type of model as in [1], in which transfer from one state to another is proportional to the number of original state, and another is SEIR type of model which includes the effect of time delay[2]. In this model, once someone gets infected, they will become contagious and later quarantined after fixed period of time^1^. Using actual num-ber of patients[5] and the activity data from mobile phone[6], we wish to know which type of model better describes the reality.

In the second section, we briefly describe the structure of two types of models, and in the third section we will see the overview of the model and fitting of the models with the real data. In the final section we will examine the result and possible future developments.

## 3 Model

Let me describe two types of models of contagion. The ordinary SEIR type of model we consider here consists of 5(or 4 for simplicity) states. *S*: susceptible(not infected), *E*: Exposed(infected but not yet contagious) *I*:Infected(infected and contagious) *Q*:Quarantined(infected but separated from others) *R*:Recovered(infected but recovered and no more contagious). Naturally the population of those 5 states adds up to the total number of population *N*. We assume that the transfer from state *S* to state *E* is proportional to *S* and *I*, because the infection happens via the contact of people in these two states. We further assume the transfer from *E* to *I* is proportional to the number of *E*(therefore it is somewhat similar to the decay of a particle to another state), and transfer from *I* to *Q* and *I* to *R* are also proportional to the number of people in state *I*.

Therefore, denoting the number of people in that particular state as the same letter, we would have a combination of equations

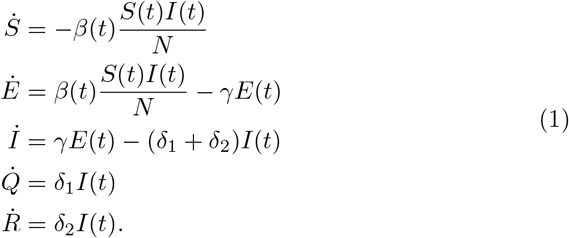

The parameter in the model can be estimated by following clinical consideration; Since it is known one will be infectious after 3 to 5 days after the infection, the time scale for transferring from E to I can be estimated to be around 4^2^. Time scale for transfer from *I* to *Q* can be estimated to be around 1 week considering the situation in Japan[4], but expected to vary from country to country. The transfer parameter *δ*_2_ from state *I* to state *R* is expected to be around 1/14, because it is considered it takes about two weeks for COVID-19 to be cured(to become not infectious). Another type of model is SEIR type of model with delay in time and the equations are given as below.

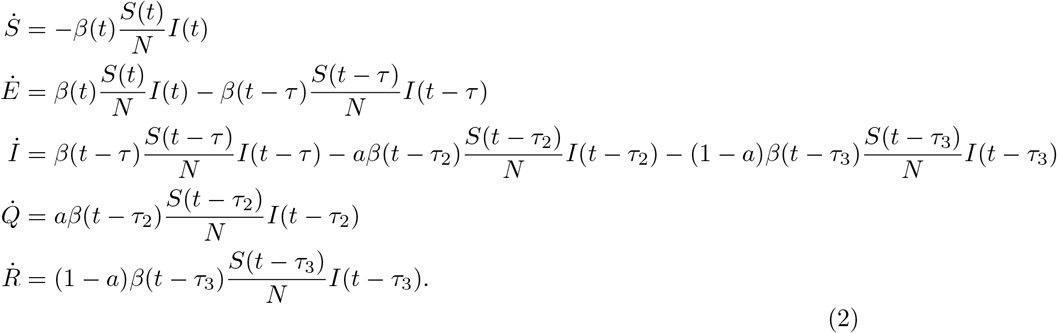

In reality, once you get infected with coronavirus, becoming symptomatic and being tested positive happens after certain amount of time rather than happening at certain probability. Thus, we have a heuristic reason to believe the latter model better describes the reality. In the later discussion in this note, we will focus on the delayed SEIR model and leave the comparison between the two models for appendix. If transfer from *S* to *E* happens at time t, transfer from state *E* to state *I* happens at time *t* + *τ*, where *τ* ∼4, and assuming that state *I* will proceed to state *Q* with probability *a* and to state R with probability (1 − *a*), we further assume that the transfer from state *I* to state *Q* will happen a t time *t* − *τ*_2_ and *I* to *R* at time *t* − *τ*_3_.

## 4 Data and anaysis

After brief description on the datasets, we will perform three different data analysis to know which of the two models above describe the dynamics of confirmed cases better including the effect of lockdown.

### 4.1 Data

We use the data for the confirmed cases from ourworlddata.org[5] and mobility data from apple mobility report[6]. The apple mobility report records how many times people searched for certain path using apple map, and the number of search is counted for car, walk and transit respectively.

### 4.2 First Analysis

In the first analysis, we wish to know the time lag from infection to quarantine (*τ*_2_) heuristically. If the value of *β* and *S* is constant, the growth rate^3^ of *I* should be proportional to *β*. Therefore, naively, we could expect that the growth rate of *Q* should be the same, so we might expect that the value of *β* and growth rate of 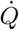 (= daily confirmed cases) is proportional. We have changed the value of *τ*_2_ and performed the linear regression of growth rate of 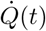 with respect to *β*(*t* −*τ*_2_), and computed p-value for each regression.

There are roughly 5 countries which has already achieved the drastic decrease in the number of confirmed cases and data is available, Australia, Austria, Czech, Norway and Switzerland. We would focus on those 5 countries. Among those, please take a look at figures for Australia and Austria below. First figure shows the number of confirmed cases, and the second shows the growth rate of the number of confirmed cases and activity index taken by [6]^4^. In the third figure, growth rate in confirmed cases and activity 12 days before was plotted.

You can see that the growth rate changes in response to the change of activity index with around 10 days of delay. We have also performed linear regression on growth rate of confirmed cases and activity index, changing the time lag *τ*. As a result, we will have p-value for the regression and we have plotted the log(p-value) in the Figure 8. If we interpret the location of minimum p-value as the most probable model^5^, we can conclude *τ* ∼ 12.

**Figure 1:**
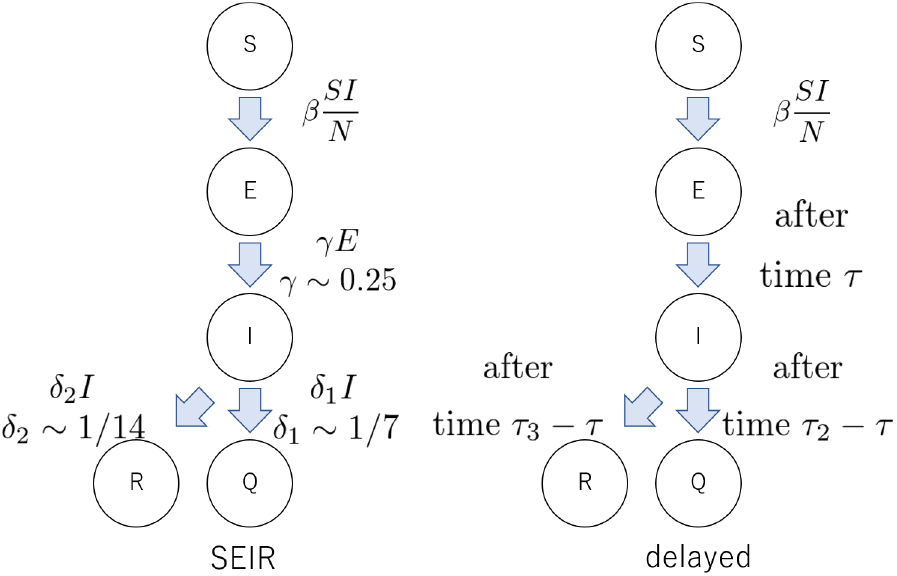
Transfer between states in SEIR model and delayed model.

**Figure 2:**
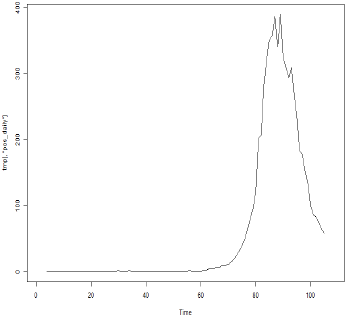
daily confirmed cases in Australia.

**Figure 3:**
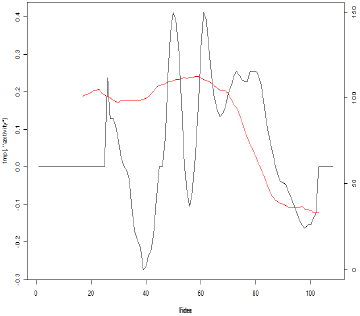
activity index and growth rate of confirmed cases in Australia.

**Figure 4:**
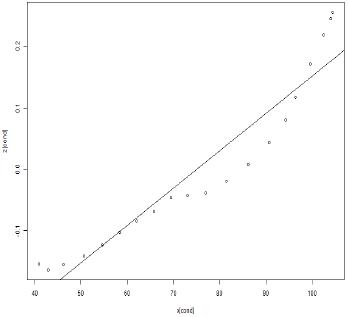
activity index(12 days before) and growth rate of confirmed cases in Australia.

**Figure 5:**
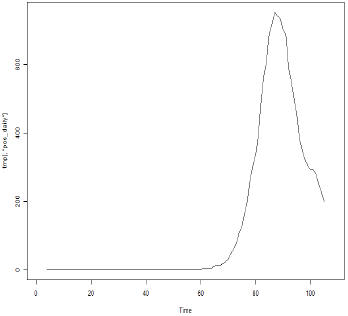
daily confirmed cases in Austria.

**Figure 6:**
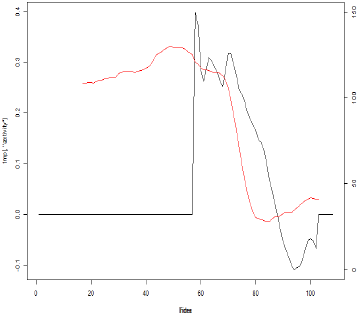
activity index and growth rate of confirmed cases in Austria.

**Figure 7:**
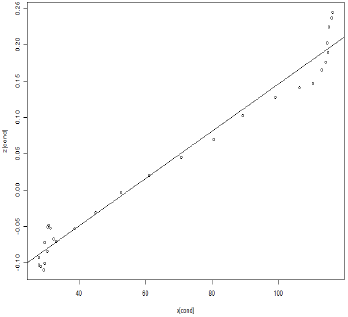
activity index(12 days before) and growth rate of confirmed cases in Austria.

**Figure 8:**
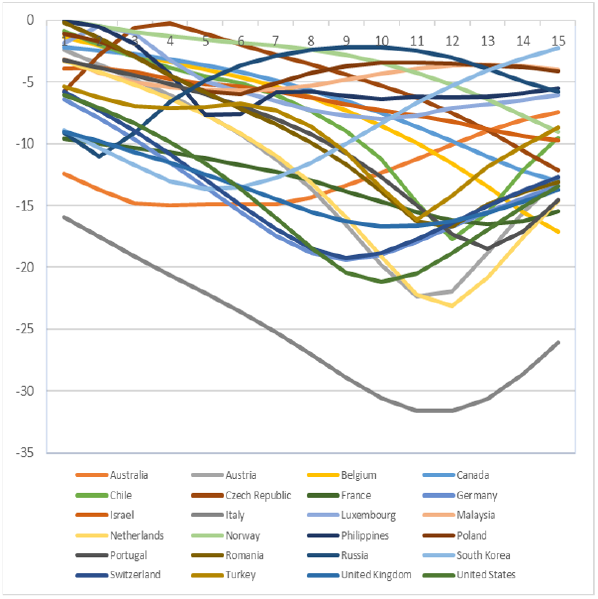
log(p-value) of linear regression of growth rate of confirmed cases with respect to activity index, changing time lag *τ*.

### 4.3 Second Analysis

In the next step, let’s perform two types of simulation I have described, SEIR type model and SEIR type model with delay. In this part, we simplify the analysis for ease of analysis. Here we ignore the state *R* because it is a bit hard to know the ratio of patients who will not quarantined but be cured without any treatment.

For the former model, we have variables the number of people for *S,E,I,Q*, and total number of people *N*, and transfer coefficients *γ, δ* are given by medical consideration. We will fit the model to the data by tuning *I*_0_(*I* at certain time), and the value of *β*. We naively expect the value of *β* to be proportional to activity index, but then it will not match the observation. We then assume that the time dependence of *β* is similar to that of activity index. That is, as in Figure 9, we first approximate the dynamics of activity index as constant for *t* < *t*_1_ and *t* > *t*_2_, and linearly decrease for *t*_1_ < *t*≦ *t*_2_. We then assume that *β* is constant for *t* < *t*_1_ and *t* > *t*_2_, and linearly decrease for *t*_1_ < *t*≦ *t*_2_.

**Figure 9:**
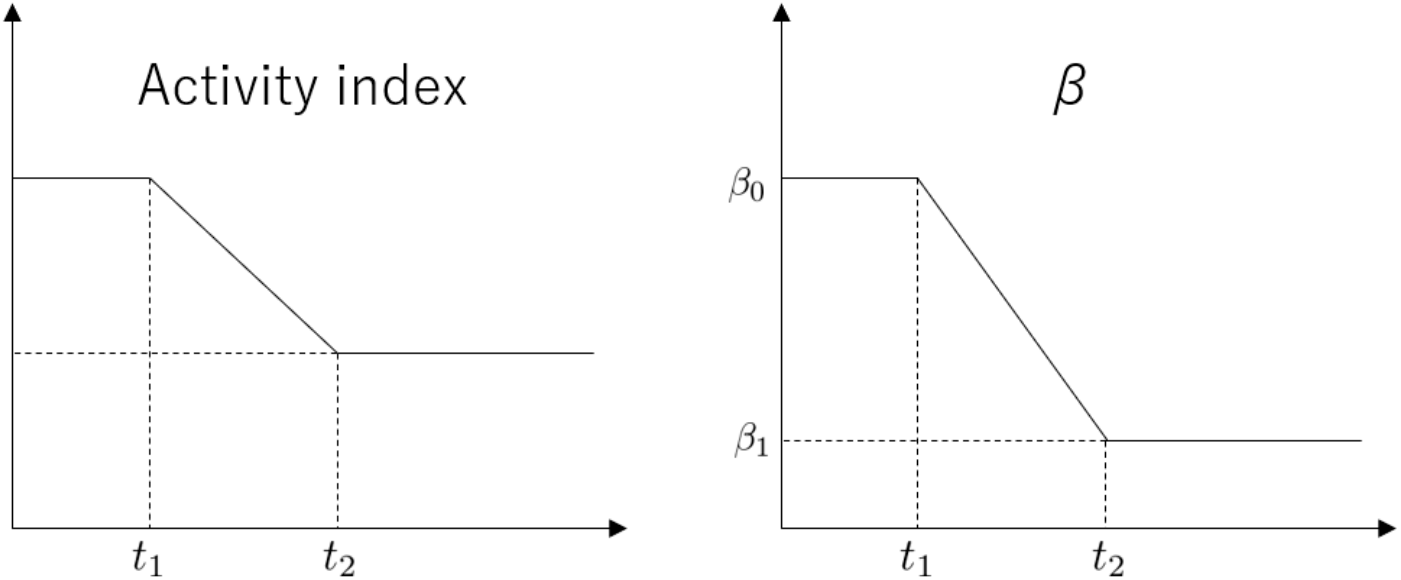
Time dependence of activity and the value of *β*.

In the delayed SEIR model, we instead have *N* and *τ* from clinical consideration, because it is known that the time period from infection till one gets infectious is considered to be around 4 days. We then fit the model to daily confirmed cases and obtain *τ*_2_, *β*_0_, *β*_1_, *I*_0_.

If we wish to see 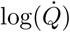, *I*_0_ corresponds to the intercept of the line, *β*_0_ corresponds to the slope of the line, *τ*_2_ corresponds to the time of lockdown, and *β*_1_ corresponds to the slope of the line after lockdown, so it is not so hard to estimate the value of the parameters. Here we adjust those parameters by hand and saw the value of 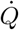 for actual and model. Here we show plot of those together with the plot of *β*.

For Australia, the parameter for the delayed model is,*β*_0_ = 1.0, *β*_1_ = 0.04*β*_0_, *τ* = 4, *τ*_2_ = 12, *I*_0_ = 0.01, *t*_0_ = 2/2, *t*_1_ = 3/12, *t*_2_ = 3/23. For Austria, the parameter for the delayed model is *β*_0_ = 0.7, *β*_1_ = 0.07*β*_0_, *τ* = 4, *τ*_2_ = 14, *I*_0_ = 6, *t*_0_ = 3/1, *t*_1_ = 3/7, *t*_2_ = 3/15. Here *t*_0_ denotes the time when the number of confirmed cases became more than or equal to 10 for the first time (and we set the day to t=0 when we draw the graph for the number of patients). Considering the fact that the value of the activity became 20% of that due to lockdown in Australia as in Figure 10, and 30% in Austria as in Figure 13, the decline in *β* in Figure 11, Figure 14 were much more drastic compared to the decline in the value of the activity. From the fact that the ratio of *β*_1_ to *β*_0_ is close to the square of ratio of activity index, we can guess that the value of *β* might just be proportional to square of activity index^6^. SEIR model with time delay managed to reproduce the real data as in Figure 12, and Figure 15. Please refer to the appendix for the data fitting for SEIR model without delay.

**Figure 10:**
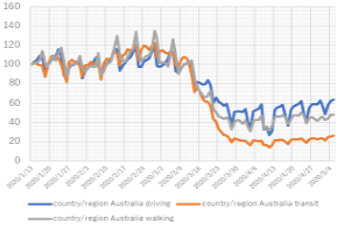
value of mobility in Australia.

**Figure 11:**
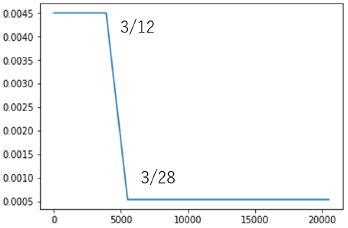
value of *β* in Australia.

**Figure 12:**
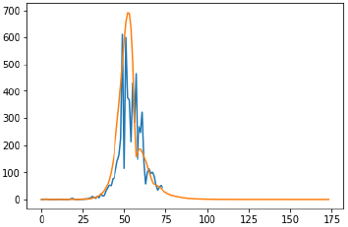
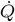 for actual and delayed model in Australia.

**Figure 13:**
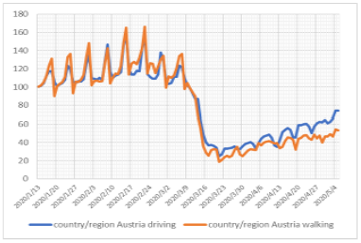
value of mobility in Austria.

**Figure 14:**
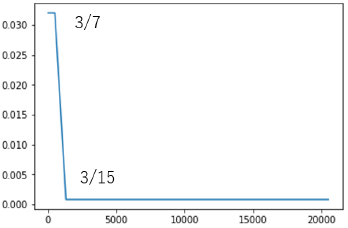
value of *β* in Austria.

**Figure 15:**
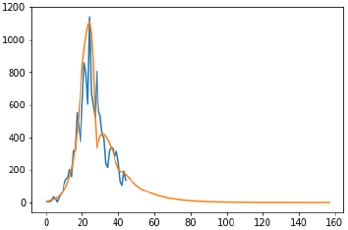
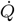 for actual and delayed model in Austria.

**Figure 16:**
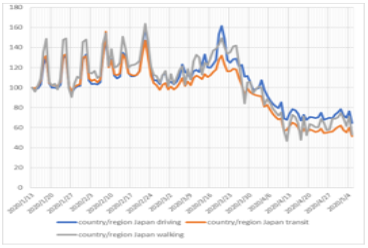
value of mobility in Japan.

**Figure 17:**
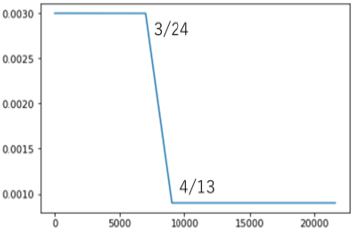
value of *β* in Japan.

**Figure 18:**
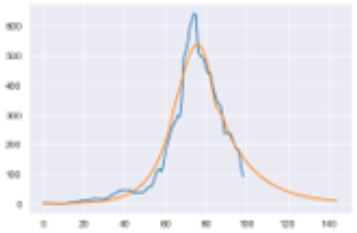
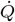 for actual and delayed model in Japan.

**Figure 19:**
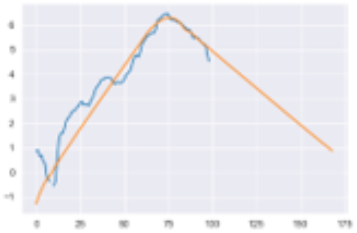
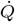 for actual and delayed model in Japan (in log scale)

**Figure 20:**
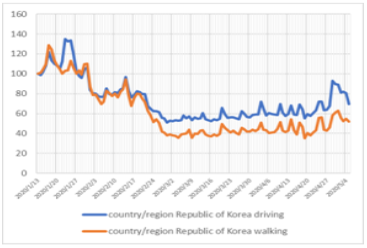
value of mobility in South Korea.

**Figure 21:**
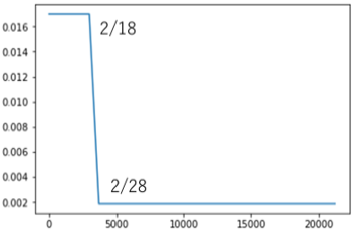
value of *β* in South Korea.

**Figure 22:**
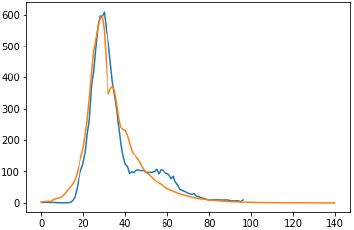
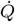 for actual and delayed model in South Korea.

**Figure 23:**
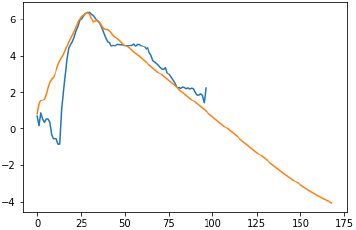
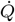 for actual and delayed model in South Korea (in log scale)

**Figure 24:**
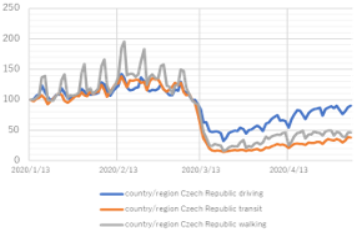
value of mobility in Czech Republic.

**Figure 25:**
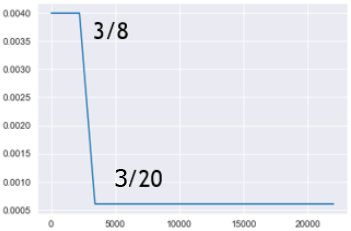
value of *β* in Czech Republic.

**Figure 26:**
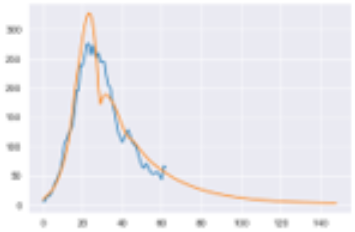
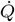 for actual and delayed model in Czech Republic.

**Figure 27:**
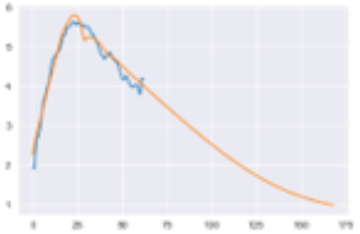
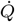 for actual and delayed model in Czech Republic (in log scale)

**Figure 28:**
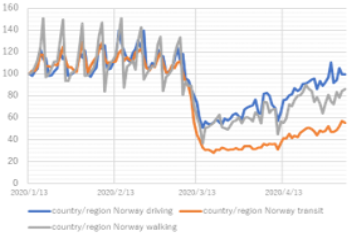
value of mobility in Norway.

**Figure 29:**
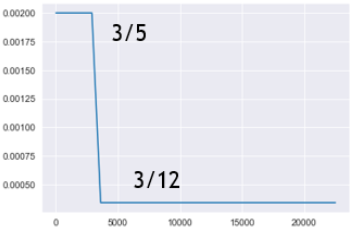
value of *β* in Norway.

**Figure 30:**
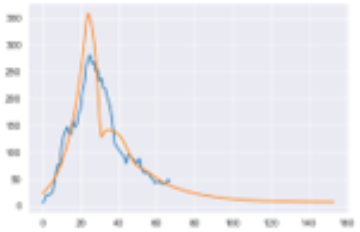
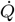 for actual and delayed model in Norway.

Let’s try to fit the delayed model for the rest of the countries which experienced successful lockdown. With respect to each country, the estimated parameters would be as in table 1. The error between real value and theoretical value from delayed SEIR model is measured by 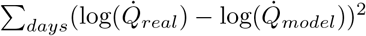.

**Table 1:** The parameters when we fit delayed SEIR model to the data.

| country | $\beta_0$ | $\beta_1/\beta_0$ | $\tau$ | $\tau_2$ | $I_0$ | $t_0$ | $t_1$ | $t_2$ | $t_2 - t_1$ | error |
| --- | --- | --- | --- | --- | --- | --- | --- | --- | --- | --- |
| Japan | 0.3 | 0.3 | 4 | 11 | 0.8 | 1/30 | 3/24 | 4/13 | 20 | 66.9 |
| South Korea | 1.0 | 0.2 | 4 | 7.2 | 1.5 | 2/1 | 2/19 | 2/26 | 7 | 35.3 |
| Czech Republic | 0.4 | 0.15 | 4 | 15 | 20 | 3/6 | 3/8 | 3/20 | 12 | 4.6 |
| Norway | 0.2 | 0.17 | 4 | 20 | 100 | 3/1 | 3/5 | 3/12 | 7 | 16.3 |
| Switzerland | 0.6 | 0.1 | 4 | 13 | 10 | 2/29 | 3/8 | 3/19 | 11 | 4.1 |
| Singapore | 0.29 | 0.17 | 4 | 16 | 0.04 | 1/20 | 3/19 | 4/10 | 22 | 649.4 |
| United States | 1 | 0.1 | 4 | 13.7 | 0.004 | 2/3 | 3/8 | 3/22 | 14 | 334.0 |
| Luxembourg | 1.1 | 0.12 | 4 | 9 | 15 | 3/13 | 3/5 | 3/20 | 15 | 3.0 |
| Vietnam | 1.2 | 0.15 | 4 | 5.5 | 0.08 | 2/5 | 3/3 | 4/1 | 29 | 83.7 |

The change of *β* and the resulting plot for confirmed cases per day will be as in Figure 16 ∼Figure 51. The ratios of activity index before and after the lockdown are respectively roughly 30% for Czech, 40 ∼ 50% for Norway, and 40% for Switzerland, and square of those numbers roughly correspond to the ratio of *β* before and after lockdown, again for those countries. ^7^

**Figure 31:**
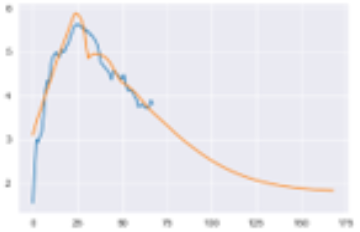
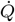 for actual and delayed model in Norway (in log scale)

**Figure 32:**
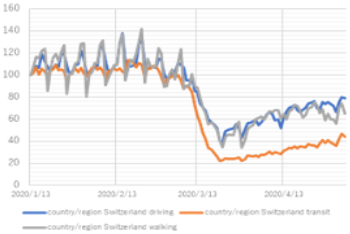
value of mobility in Switzerland.

**Figure 33:**
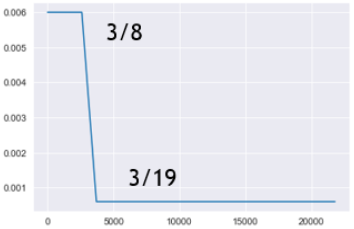
value of *β* in Switzerland.

**Figure 34:**
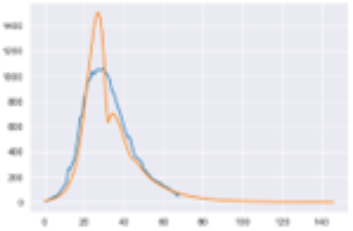
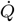 for actual and delayed model in Switzerland.

**Figure 35:**
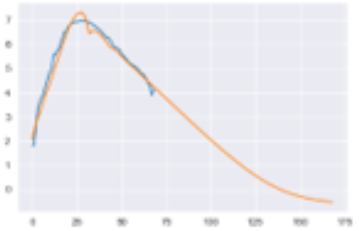
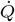 for actual and delayed model in Switzerland (in log scale)

**Figure 36:**
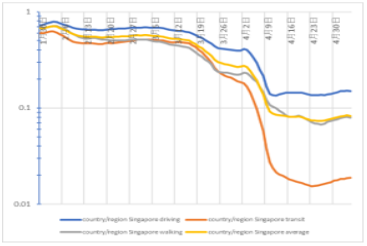
value of mobility in Singapore.

**Figure 37:**
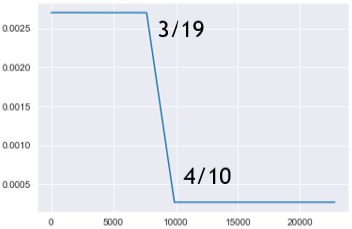
value of *β* in Singapore.

**Figure 38:**
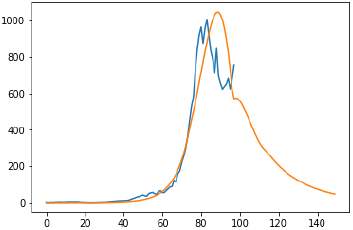
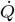 for actual and delayed model in Singapore.

**Figure 39:**
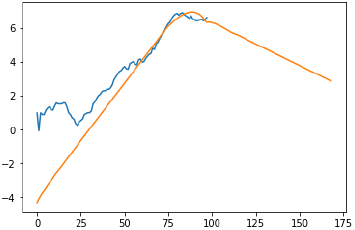
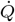 for actual and delayed model in Singapore (in log scale)

**Figure 40:**
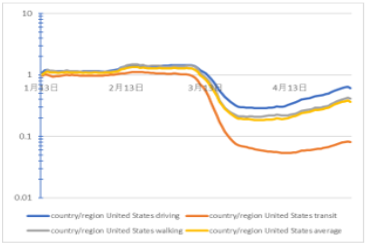
value of mobility in United States.

**Figure 41:**
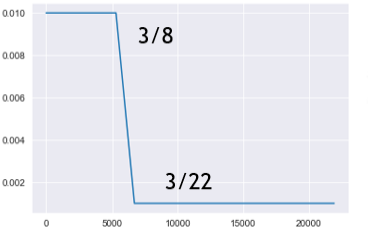
value of *β* in United States.

**Figure 42:**
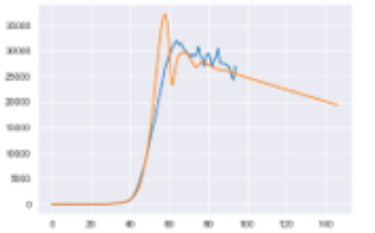
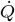 for actual and delayed model in United States.

**Figure 43:**
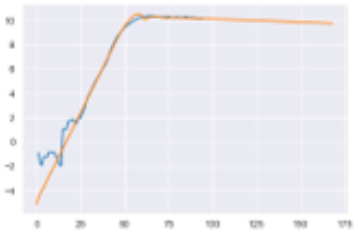
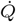 for actual and delayed model in United States (in log scale)

**Figure 44:**
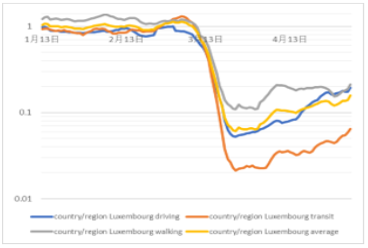
value of mobility in Luxembourg.

**Figure 45:**
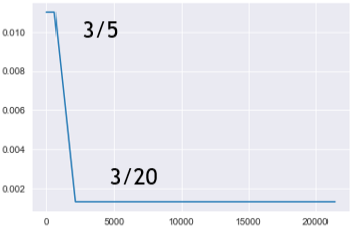
value of *β* in Luxembourg.

**Figure 46:**
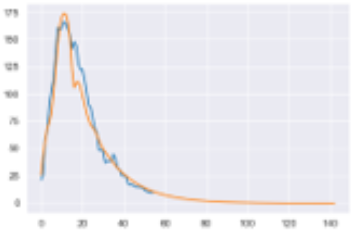
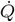 for actual and delayed model in Luxembourg.

**Figure 47:**
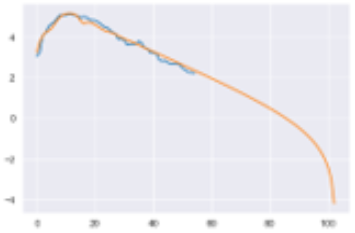
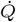 for actual and delayed model in Luxembourg (in log scale)

**Figure 48:**
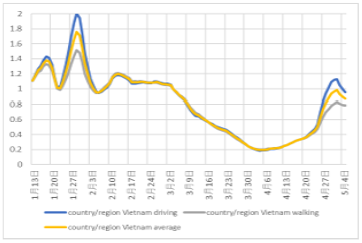
value of mobility in Vietnam.

**Figure 49:**
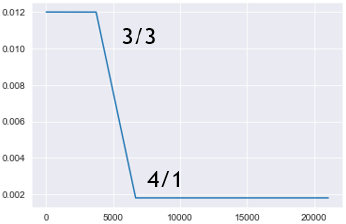
value of *β* in Vietnam.

**Figure 50:**
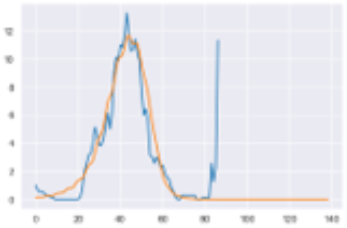
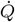 for actual and delayed model in Vietnam.

**Figure 51:**
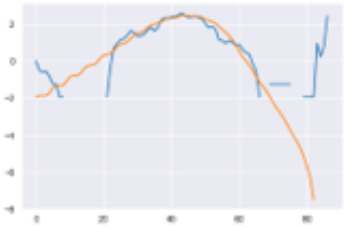
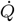 for actual and delayed model in Vietnam (in log scale)

### 4.4 Third Analysis

We will improve the analysis in the previous subsection in two ways; Firstly, we have seen that the contact coefficient *β* is roughly proportional to (*mobility*)^2^, and then we reverse the logic and assume this proportionality relationship to derive the value of *β*. We will again use [6], and use two possible mobility indices, one is average of mobility indices of three transportation(driving, transit, walking)^8^, and the other is mobility index for transit. Secondly, in the previous subsection, we have assumed fixed period of time one requires from the time of infection till the period one gets infectious, *τ*, and from the time of infection till the period one gets tested and quarantined, *τ*_2_. Here instead we assume Weibull distribution for both time lag. From the analysis of [3], the distribution of the time period from the infection to starting to be symptomatic is, Weibull distribution with mean 6.4 and standard deviation 2.3. Considering the fact one becomes infectious 2 ~ 3 days before one becomes symptomatic[8], we could pretend that the distribution of the time period from the infection to starting to be infectious is Weibull distribution with mean 4 and standard deviation 2.3. This amounts to Weibull distribution with parameter *k* ∼1.8 and *λ* ∼ 4.5^9^. Furthermore, in case of Japan, the distribution from the time of becoming symptomatic and the time of getting tested is known [9], and adding typical time from infection till one gets tested, which is 6.4 days, to this distribution, the distribution of time from infection till getting tested can be approximated as another Weibull distribution, with *k* = 3.3 and *λ* = 13.3. We assume that for the corresponding distribution in other countries, the value of *k* is 3, and fit the *λ* parameter for this distribution(denote as *λ*_2_). As a result, fitting the curve by using *β* = (mobility of public transportation)^2^ seems to be the best fit, so we will show the fit with that criterion and also show the curve from *β* = (average of three types of mobility)^2^. Moreover, we here assume *a* = 0, considering generalization to *a* = 0 will not change the dynamics qualitatively. The parameter for fitting is given as in table 2.

**Table 2:** The parameters when we fit delayed SEIR model with assumption *β* = (mobility of transit)^2^.

| country | $I_0$ | $\beta_0$ | $t_0$ | $\lambda_2$ |
| --- | --- | --- | --- | --- |
| Australia | 0.008 | 0.7 | 2/2 | 11 |
| Japan | 1.1 | 0.23 | 1/30 | 13.3 |
| Czech | 80 | 0.4 | 3/6 | 24 |
| Norway | 350 | 0.2 | 3/1 | 20 |
| Switzerland | 100 | 0.6 | 2/29 | 15 |
| Singapore | 4 | 0.35 | 1/30 | 30 |
| United States | 0.04 | 0.4 | 2/3 | 24 |
| Luxembourg | 60 | 1.3 | 3/13 | 12 |

## 5 Conclusion and Discussion

In the above discussion, as a crude data analysis, we have seen that growth rate of confirmed cases of COVID-19 is roughly linear in activity index, with time delay around 10 ± 5 days. In a more refined data analysis, we have tried to fit to the data with SEIR model and similar model with time delay and seen that it is not likely SEIR fits the real data, while the model with time delay can reproduce the data. In that analysis, we saw that the the ratio of the value of contact coefficient *β* before and after lockdown, *β*_1_/*β*_0_, is roughly equal to the ratio of square of mobility index before and after lockdown, so in the third analysis of this note, we have tried to fit the data with the assumption *β* = (mobility)^2^, using two different definition of mobility. We have confirmed that the data roughly fits this assumption, although the true value of contact coefficient should be in between the result expected by using average mobility, and the one by transit mobility. We need further investigation to know more rigorous relationship between the mobility and the contact coefficient for modelling the number of confirmed cases of COVID-19. In particular, if we wish to fit the parameters for the number of confirmed cases, we might not be able to fit well with the data for the second wave, or third wave of covid-19, so it seems we need other parameters than mobility to explain the dynamics of confirmed cases. In Japanese case, temperature could be a promising candidate for the explanatory parameter, because in the mid-late summer we saw the decrease in the number of confirmed cases.

## Data Availability

All the COVID-19 data and mobility data soruce is described in the note.

## A Appendix

In this appendix, we will see the data fitting for SEIR model without delay. Please compare this result with the delayed SEIR model we have seen above(Table 1 and Figure 12 ∼Figure 51). The curve was fitted by hand and subject to some error. The parameters for the fitting are as in Table 3. We can see that in most of the countries delayed SEIR model fits better with the same number of parameters.

**Table 3:**
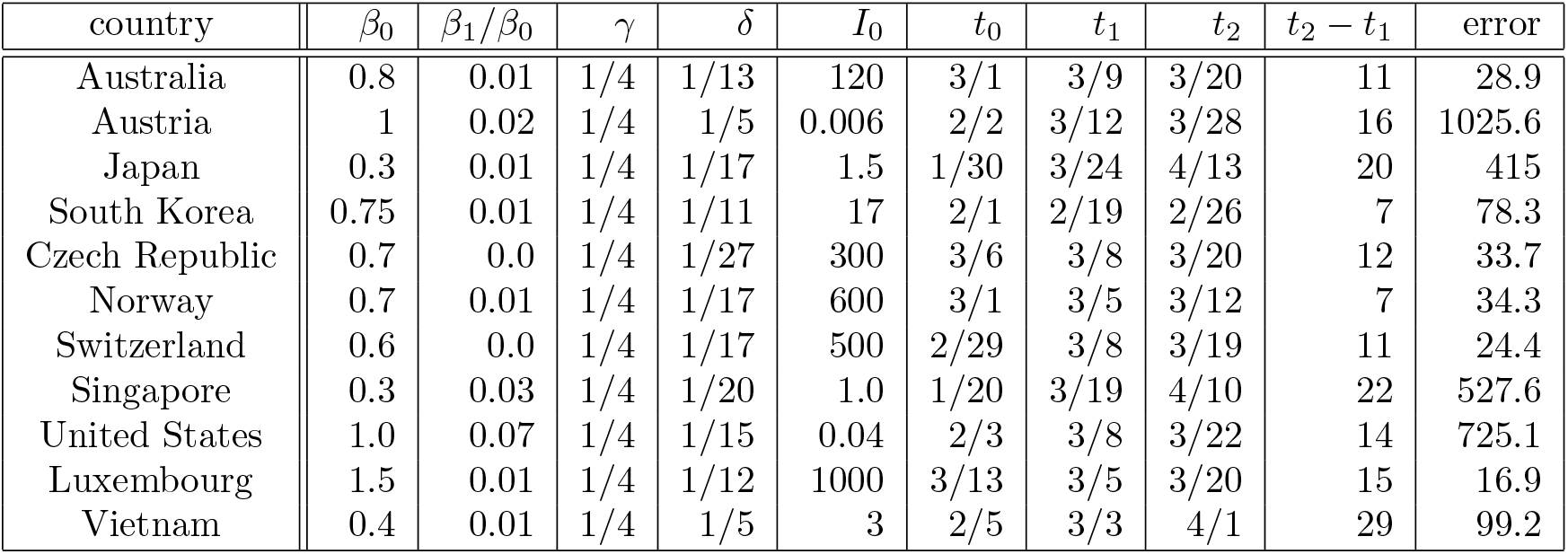
The parameters when we fit SEIR model to the data.

The figures for the fitting are as in Figure 80 ∼ Figure 101.

**Figure 52:**
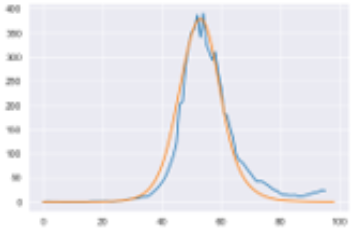
Fitting of 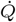 for Australia by assuming *β* = (*transit mobility*)^2^.

**Figure 53:**
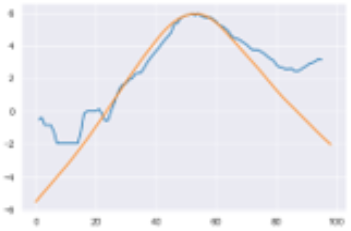
Fitting of 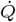 for Australia by assuming *β* = (*transit mobility*)^2^(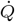 in log scale)

**Figure 54:**
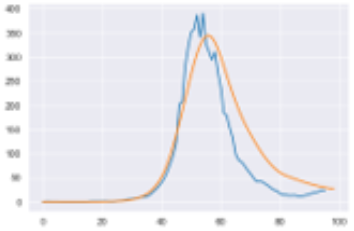
Fitting of 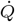 for Australia by assuming *β* = (*average mobility*)^2^.

**Figure 55:**
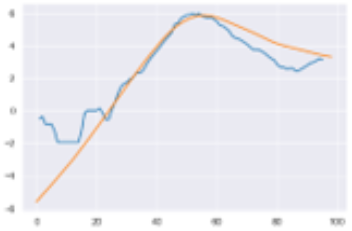
Fitting of 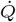 for Australia by assuming *β* = (*average mobility*)^2^(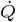 in log scale)

**Figure 56:**
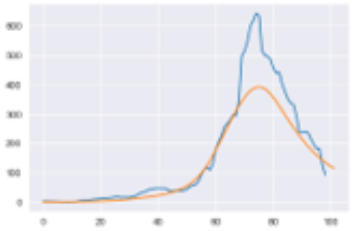
Fitting of 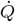 for Japan by assuming *β* = (*transit mobility*)^2^.

**Figure 57:**
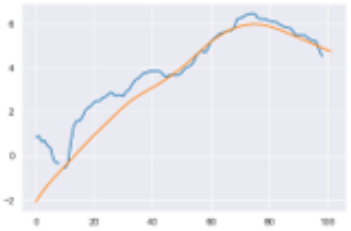
Fitting of 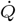 for Japan by assuming *β* = (*transit mobility*)^2^(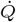 in log scale)

**Figure 58:**
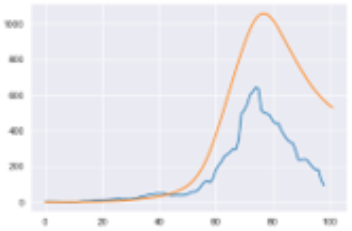
Fitting of 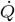 for Japan by assuming *β* = (*average mobility*)^2^.

**Figure 59:**
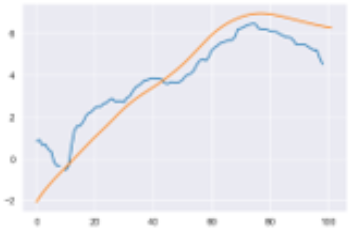
Fitting of 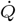 for Japan by assuming *β* = (*average mobility*)^2^(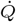 in log scale)

**Figure 60:**
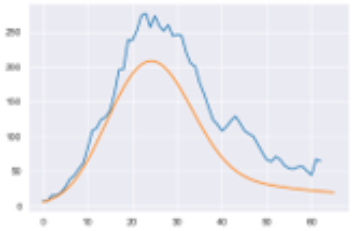
Fitting of 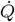 for Czech Republic by assuming *β* = (*transit mobility*)^2^.

**Figure 61:**
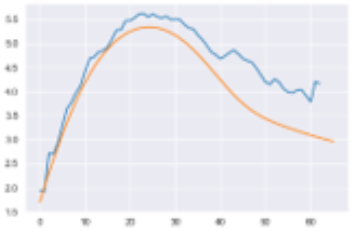
Fitting of 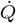 for Czech Republic by assuming *β* = (*transit mobility*)^2^(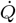 in log scale)

**Figure 62:**
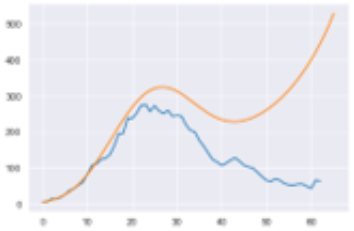
Fitting of 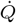 for Czech Republic by assuming *β* = (*average mobility*)^2^.

**Figure 63:**
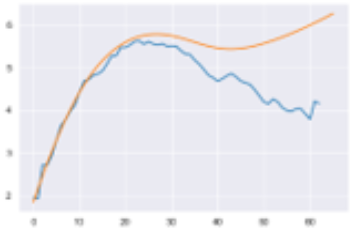
Fitting of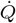 for Czech Republic by assuming *β* = (*average mobility*)^2^(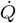 in log scale)

**Figure 64:**
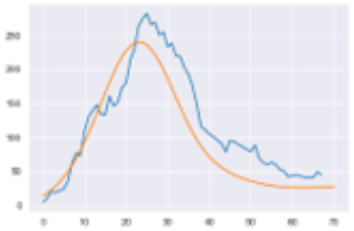
Fitting of 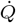for Norway by assuming *β* = (*transit mobility*)^2^.

**Figure 65:**
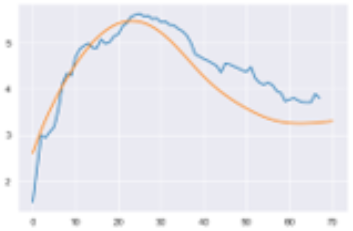
Fitting of 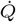 for Norway by assuming *β* = (*transit mobility*)^2^(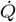 in log scale)

**Figure 66:**
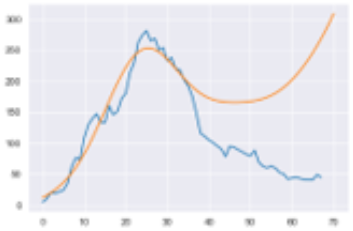
Fitting of 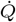 for Norway by assuming *β* = (*average mobility*)^2^.

**Figure 67:**
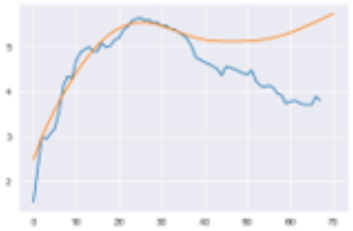
Fitting of 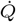 for Norway by assuming *β* = (*average mobility*)^2^(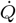 in log scale)

**Figure 68:**
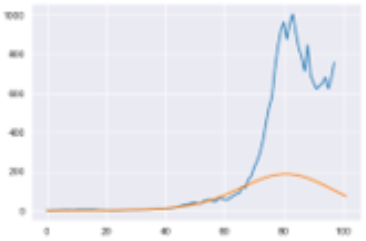
Fitting of 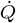 for Singapore by assuming *β* = (*transit mobility*)^2^.

**Figure 69:**
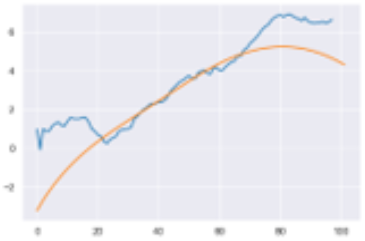
Fitting of 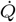 for Singapore by assuming *β* = (*transit mobility*)^2^(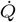 in log scale)

**Figure 70:**
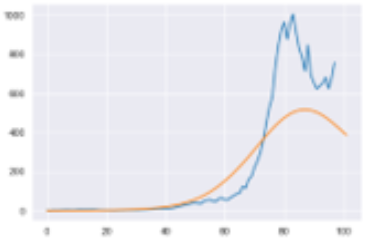
Fitting of 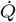 for Singapore by assuming *β* = (*average mobility*)^2^.

**Figure 71:**
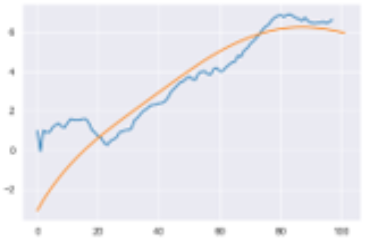
Fitting of 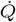 for Singapore by assuming *β* = (*average mobility*)^2^(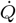 in log scale)

**Figure 72:**
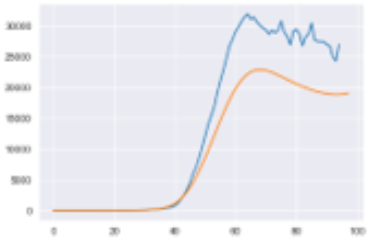
Fitting of 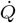 for United States by assuming *β* = (*transit mobility*)^2^.

**Figure 73:**
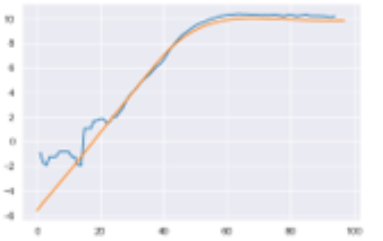
Fitting of 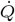 for United States by assuming *β* = (*transit mobility*)^2^(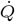 in log scale)

**Figure 74:**
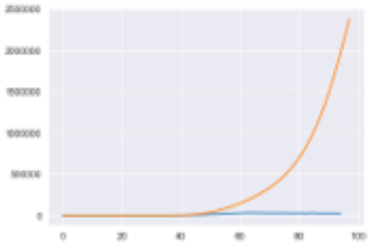
Fitting of 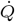 for United States by assuming *β* = (*average mobility*)^2^.

**Figure 75:**
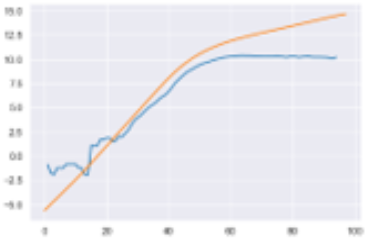
Fitting of 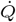 for United States by assuming *β* = (*average mobility*)^2^(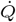 in log scale)

**Figure 76:**
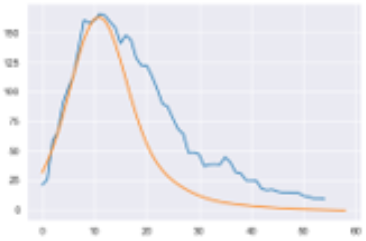
Fitting of 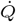 for Luxembourg by assuming *β* = (*transit mobility*)^2^.

**Figure 77:**
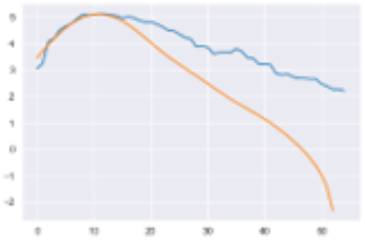
Fitting of 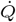 for Luxembourg by assuming *β* = (*transit mobility*)^2^(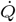 in log scale)

**Figure 78:**
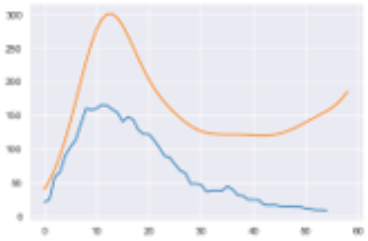
Fitting of 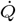 for Luxembourg by assuming *β* = (*average mobility*)^2^.

**Figure 79:**
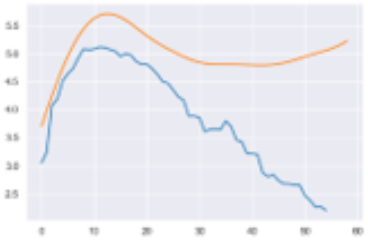
Fitting of 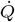 for Luxembourg by assuming *β* = (*average mobility*)^2^(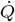 in log scale)

**Figure 80:**
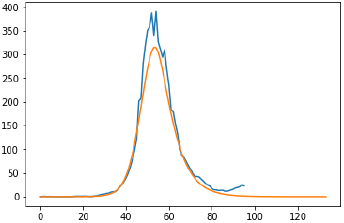
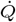 for actual and SEIR model in Australia.

**Figure 81:**
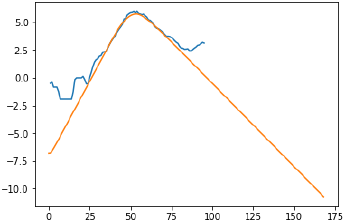
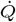 for actual and SEIR model in Australia (in log scale)

**Figure 82:**
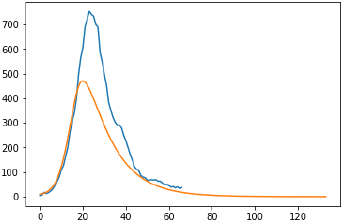
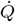 for actual and SEIR model in Austria.

**Figure 83:**
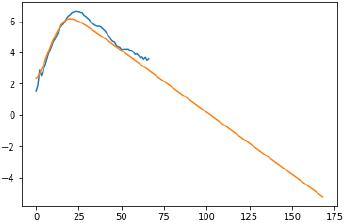
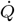 for actual and SEIR model in Austria (in log scale)

**Figure 84:**
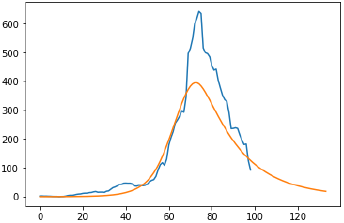
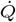 for actual and SEIR model in Japan.

**Figure 85:**
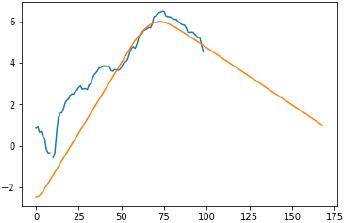
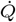 for actual and SEIR model in Japan (in log scale)

**Figure 86:**
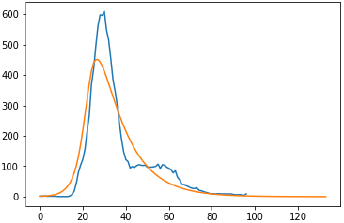
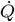 for actual and SEIR model in Korea.

**Figure 87:**
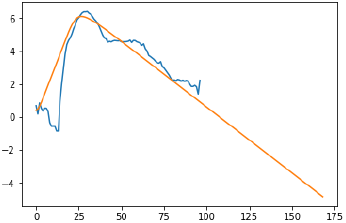
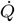 for actual and SEIR model in Korea (in log scale)

**Figure 88:**
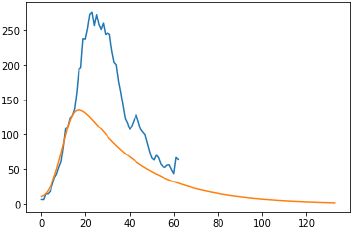
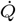 for actual and SEIR model in Czech.

**Figure 89:**
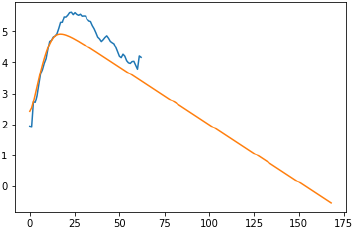
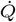 for actual and SEIR model in Czech (in log scale)

**Figure 90:**
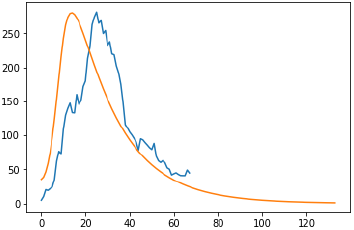
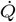 for actual and SEIR model in Norway.

**Figure 91:**
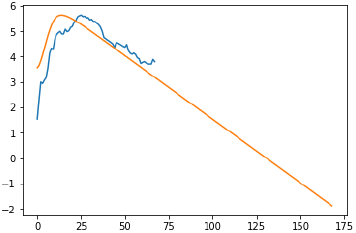
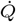 for actual and SEIR model in Norway (in log scale)

**Figure 92:**
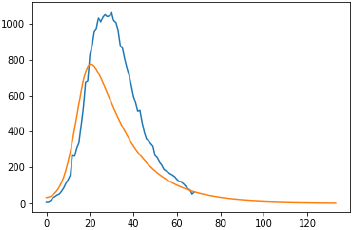
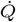 for actual and SEIR model in Switzerland.

**Figure 93:**
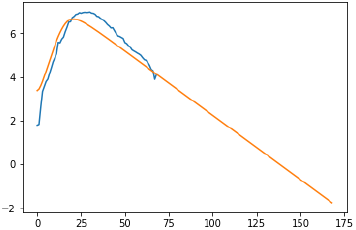
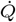 for actual and SEIR model in Switzerland (in log scale)

**Figure 94:**
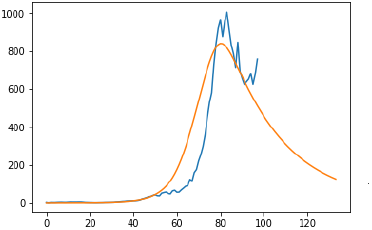
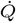 for actual and SEIR model in Singapore.

**Figure 95:**
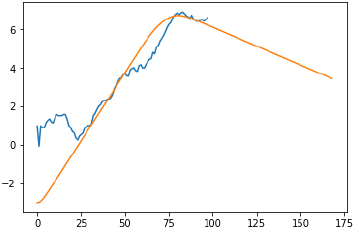
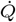for actual and SEIR model in Singapore (in log scale)

**Figure 96:**
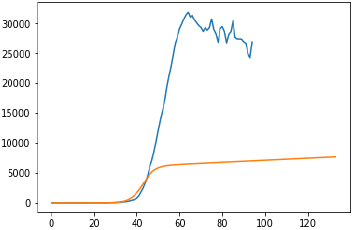
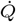 for actual and SEIR model in United States.

**Figure 97:**
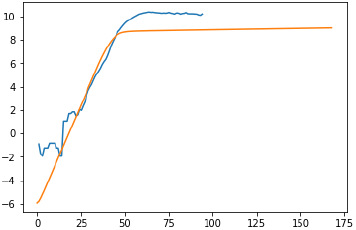
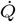 for actual and SEIR model in United States (in log scale)

**Figure 98:**
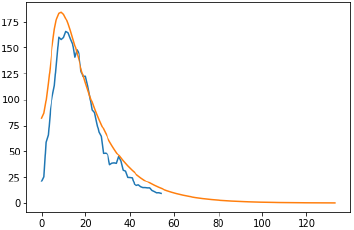
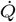 for actual and SEIR model in Luxembourg.

**Figure 99:**
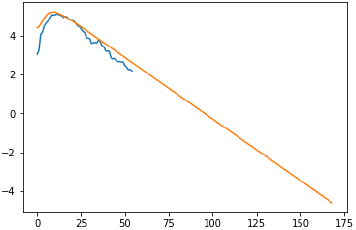
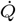 for actual and SEIR model in Luxembourg (in log scale)

**Figure 100:**
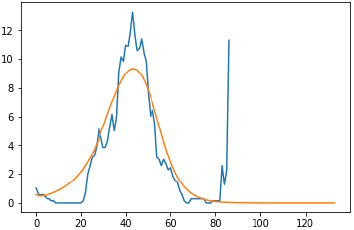
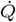 for actual and SEIR model in Vietnam.

**Figure 101:**
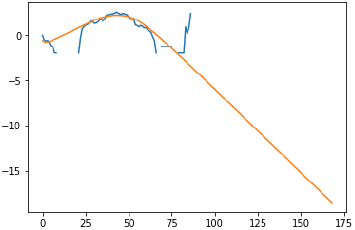
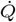 for actual and SEIR model in Vietnam (in log scale)

## Footnotes

1 In reality, we know that the time infected person becomes contagious is rather fixed[3], and also it will take a while since patients go to a doctor till they receive the result of PCR testing. Therefore we expect the model with time delay will describe the dynamics better.

2 The time from infection till one becomes mptomatic is around 5 days as in the study in Diamond Princess[3] and it is also known that one can be infectious 1 ∼ 2 days before one becomes symptomatic[8]. Therefore time scale for this process is around 4 and *γ* should be around 0.25.

3 We computed the slope of confirmed cases with respect to time as the growth rate.

4 In the original database, the amount of search people performed through apple map for walk, drive, transit are recorded and we just took the average of these indices as activity index.

5 If possible we should compare the model of different *τ* by free energy or BIC, but we don’t have time to do that.

6 That actually makes sense, because if we assume for example, only 20% of people go out and the rest stay home, both *S* and *I* will be multiplied by 20% and effectively *β* becomes square of 20%.

7 It is also claimed from purely analysing the data in Japan in some twitter accounts[7].

8 Note there is no theoretical background why we should take the average of the three values, since it is NOT weighted average of those

9 Remember that Weibull distribution is *f* (*t*) = (*k*/*η*)(*tη*)^*k*−1^ exp{−(*t*/*η*)^*k*^}

## Notes

### Competing Interest Statement

The authors have declared no competing interest.

### Funding Statement

There is no funding for this work.

### Author Declarations

I don't belong to any institution so I did not conduct any IRB/ethics check.

